# Prospective whole-genome sequencing uncovers factors influencing bacterial transmission in neonates (PROtect NEO)

**DOI:** 10.1101/2024.11.22.24317721

**Authors:** Timmy Nguyen, Fabian Bürkin, Stefany Ayala-Montaño, Iván Acevedo Monterrosa, Daniel Jonas, Daniel Klotz, Hans Fuchs, Martin Kuntz, Christian Schneider, Martin Wolkewitz, Tjibbe Donker, Sandra Reuter, Tim Götting, Philipp Henneke

## Abstract

**Importance:** Infants in neonatal intensive care units are at risk of transmission events by bacteria with multidrug-resistance and/or epidemic potential (“multidrug-resistant organisms plus”, MDRO+), which may precede invasive infections. Prospective high-level resolution of MDRO+ transmission clusters may alleviate high risk situations through targeted infection prevention control measures.

**Objectives:** 1) Exploration of whole-genome sequencing in resolving putative MDRO+ transmission chains. 2) Analysis of risk factors for becoming part of a transmission cluster.

**Design, Setting, Participants:** Prospective monocentric cohort study at a level III neonatal intensive care unit at the Medical Center – University of Freiburg, Germany. Inclusion of 434 of 551 preterm and term infants admitted for at least 48h and screened at least once between February 15 2019 and November 16 2020.

**Exposures:** Integration of (1) culture-based screening, (2) genetic typing with amplified fragment length polymorphism and whole-genome sequencing and (3) granular clinical and staffing data. Statistical analysis of time-dependent risk factors based on advanced multivariate model analysis.

**Main Outcomes:** Primary: Identification of MDRO+ transmission events, indistinguishable by amplified fragment length polymorphism or whole-genome sequencing. Secondary: MDRO+ colonization rates; identification of factors influencing transmission events; MDRO+ blood stream infection rates.

**Results:** Among 434 participants 51.8 % (95% CI, 47.1%-56.5%) were colonized with at least one MDRO+; 32.5% (95% CI, 28.3%-37.0%) were colonized by transmission. Among 38 unique transmission clusters, *E. coli* was the most common cluster-forming MDRO+. Four of ten MDRO+ blood stream infections originated from transmission events. Multivariate analysis revealed three factors influencing the risk of becoming part of a transmission cluster: Increased nurse staffing levels and antibiotic administration lowered the risk of becoming part of a bacterial transmission cluster, while vascular catheter usage increased it.

**Conclusions and Relevance:** Prospective whole-genome sequencing of routine screening isolates from neonatal intensive care unit infants is powerful for detecting MDRO+ transmission chains, exceeding amplified fragment length polymorphism in precision and seems justified in high-risk neonates to uncover specific risk factors for MDRO+ transmission. Delayed and “false” identification of transmission events, which inevitably occur in conventional microbiological screening, have grave organizational consequences.

## Introduction

In healthy infants, the establishment of metabolic independence after birth is associated with acquisition of maternal microorganisms from birth canal, skin and breast milk. This drives the dynamic microbiome development in a postnatal relationship between mother and infant best termed ‘separate, but intertwined’. However, in hospitalized and in particular in preterm infants, the microbiome receives major input from hospital-adapted microorganisms residing in other patients and on inanimate surfaces. Conceptually, these microorganisms are qualitatively distinct, given that they have evolved traits shaped by hosts that carry diseases, by antibiotic selection pressure and by the hospital environment. In contrast to older children and adults, the microbiome of infants is less stable, i.e. less resilient to the incorporation of new strains. The altered antimicrobial resistance in neonates acutely links microbiome composition and thus hospital-adapted bacteria to life-threatening individual infections and infection outbreaks on Neonatal Intensive Care Units (NICUs). We recently confirmed the difficulty of preventing transmission events (TEs) in infants even with stringent hygiene measures in a randomized probiotic study: Over 48 percent of infants in the placebo group, i.e. without direct contact with the probiotics, eventually carried the probiotic *Bifidobacterium* strain [1].

Overall, the gradual colonization of infants in NICUs includes bacteria with multidrug resistance (MDR) or increased potential for patient-to-patient transmission [2–5]. Nosocomial bacterial colonization, antibiotic usage and indwelling catheters increase the risk of nosocomial infections (NI) [6] and death [7]. Factors influencing bacterial colonization include prematurity, mode of delivery [8,9], antibiotic therapy [10,11] and the hospital environment [12]. In contrast, factors driving TEs in NICUs remain largely elusive. The spatial and temporal clustering of bacteria of the same species is strongly suggestive of an outbreak [13] and should be investigated with appropriate typing methods to confirm or refute putative TEs [14]. Despite extensive infection prevention control (IPC) measures, including environmental investigations, point sources are infrequently discovered [15,16], indicating that TEs predominantly occur via indirect contact, e.g. the skin of health care personnel. In Germany, weekly microbiological surveillance has been recommended for very low birthweight infants (VLBW, < 1500 g birth weight) by the Robert Koch-Institute (RKI) since 2012 [17]. This culture-based screening targets potentially problematic bacterial species, i.e. species belonging to the order of *Enterobacterales* and non-fermenters declared MDR Gram-negative bacteria, and methicillin resistant *Staphylococcus aureus* (MRSA) [18], together with their antibiotic sensitive counterparts, which are collectively referred to as “multidrug resistant organisms plus” (MDRO+) in this study [1]. MDR Gram-negative bacteria are relatively rare in NICUs in Germany and many other European countries, i.e. their parallel, species-identical detection in multiple patients is highly indicative of a TE. However, this is not the case for parallel detection of the more frequent non-MDR *Enterobacterales*, since they often represent distinct strains of the same species, yet with similar resistance patterns. Accordingly, the extent of recommended and potentially disruptive organizational IPC measures, ranging from cohorting of patients and staff to environmental sampling and ward closures, is often at odds with inaccurate MDRO+ characterization, in particular their genomic relationship. Moreover, a stepwise complex typing of isolates usually introduces considerable delay, so that the necessity of the hygiene measures taken can only be assessed retrospectively.

Here we asked how timely and accurate TE characterization by amplified fragment length polymorphism (AFLP) and whole genome sequencing (WGS) impacts the management of transmission clusters (TC) and how, at the proposed granular level, treatment and management strategies influence the emergence and development of MDRO+ clusters in the NICU.

## Methods and Analysis

### Design, Setting, Participants

This single-center prospective cohort study was conducted at a 17-bed Level III NICU (Medical Center – University of Freiburg, Germany), managing preterm and term newborns. 434 newborns were enrolled from February 15 2019, to November 16 2020. Patients admitted to the NICU for ≥ 48 hours with ≥ 1 screening were included; those with shorter stay or no screening were excluded. *Figure 1* outlines the study design. The initial protocol was published beforehand [19]. In accordance with the RKI recommendations [17], culture-based screening was performed on admission and on each subsequent Sunday.

**Figure 1:**
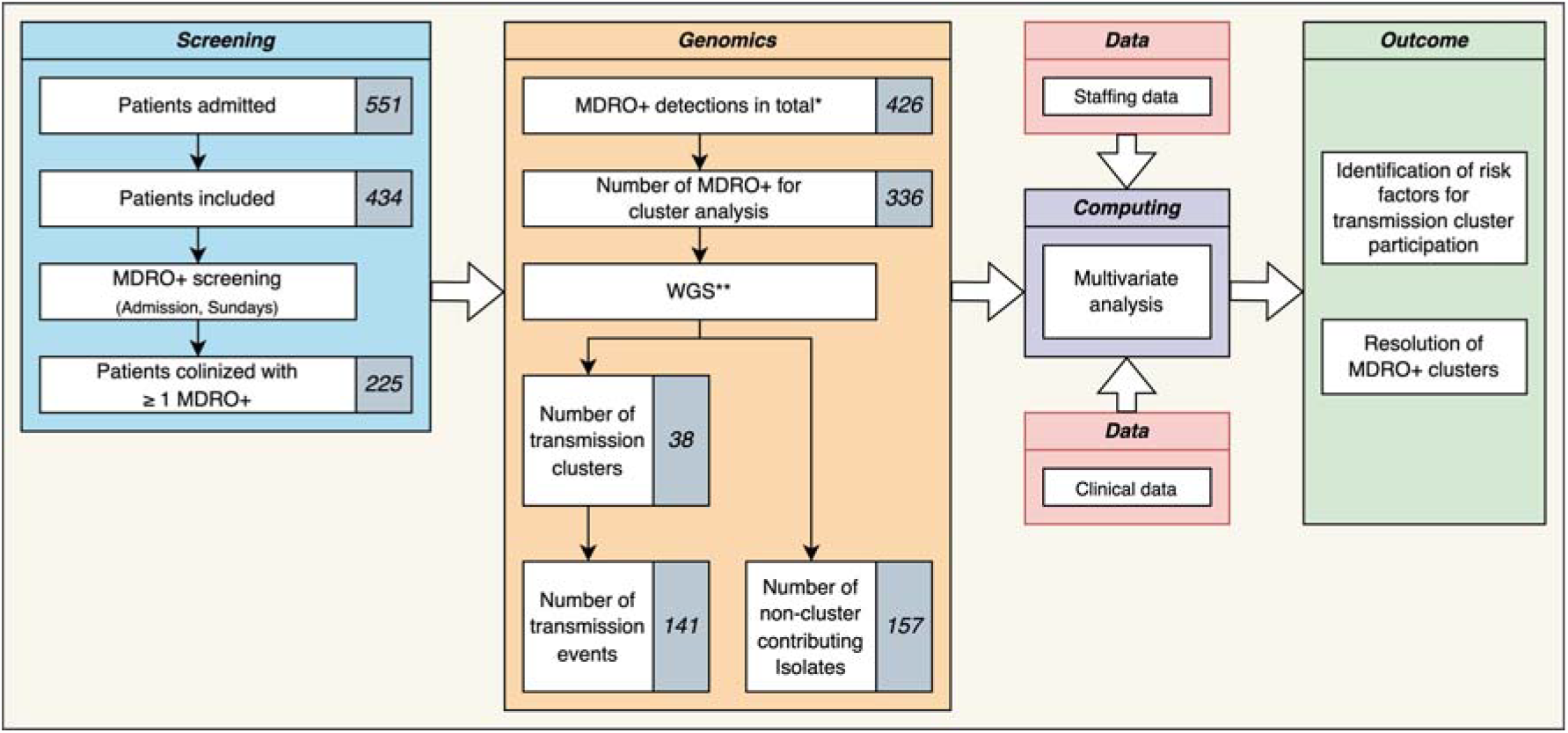
Study Outline Flow Chart. From the included patients, bacterial isolates (MDRO+) from weekly screenings were used to determine genomic clusters using WGS. The genomic information was then combined with clinical and staffing data for multivariate logistic regression analysis. The genomic analysis resolved putative MDRO+ clusters, and multivariate analysis identified associated influencing factors that either increased or decreased an infant’s risk of becoming part of a transmission cluster within the boundaries of the model. * The total number of MDRO+ detections was independent of patient allocation ** AFLP typing was performed as well, yet final cluster determination was based on WGS

### Outcomes

#### Primary outcome

Identification of bacterial TEs defined as indistinguishable same-species-pathogens in different patients after molecular/genomic typing.

#### Secondary outcomes

Rate of patients colonized with ≥ 1 MDRO+, rate of MDRO+ colonization and rate of blood stream infections per 1,000 patient days. Factors influencing the risk of TC participation.

#### Bacterial Transmission – Definitions according to WGS

**Transmission cluster (TC)**: ≥ 2 genetically indistinguishable bacterial isolates from different patients, including the index patient.

**Transmission event (TE)**: Instance where a bacterial isolate is transmitted between patients.

#### Collection of Data

Data collected:

- Patient and microbiological diagnostic data
- Anonymized G-BA (German: “Gemeinsamer Bundesausschuss”) nurse staffing data [20]

Bacterial screening isolates were transferred from the microbiology department to the typing laboratory. Swabs sites included the nasopharynx, the anorectal junction and other risk sites (e.g. stomata). For further information see supplement (*<u>eMethod 2, 3, 4</u>*).

#### Genetic Typing of Bacterial Strains

AFLP-typing for gram-negative bacteria, *spa*-typing for Methicillin susceptible *S. aureus* (MSSA) and MRSA, using Genetic Analyzer abi3500 and analytical software *GeneMapper*™ (Thermo Fisher Scientific) or *BioNumerics* (Applied Maths NV). WGS was based on the *MiSeq Nextera* platform (Illumina). Data is stored in ENA project PRJEB81699. For further information on genetic typing and accessions see supplement (*<u>eMethod 5</u>*).

#### Statistical Analysis

Data analysis was performed with *Excel* (Microsoft) and *R* (v4.4.1). Outliers were retained. Following Barnett & Graves [21] and Breslow et al. [22], temporal dynamics of the multivariate risk factor analysis for TEs was mapped using a logistic regression model, which allows a differentiated view of relevant time periods before a TE could be detected. Temporal relationships between TE and influencing variables X_i_ are shown in *Figure 2*. For further information see supplement (*<u>eMethod 6</u>*).

**Figure 2:**
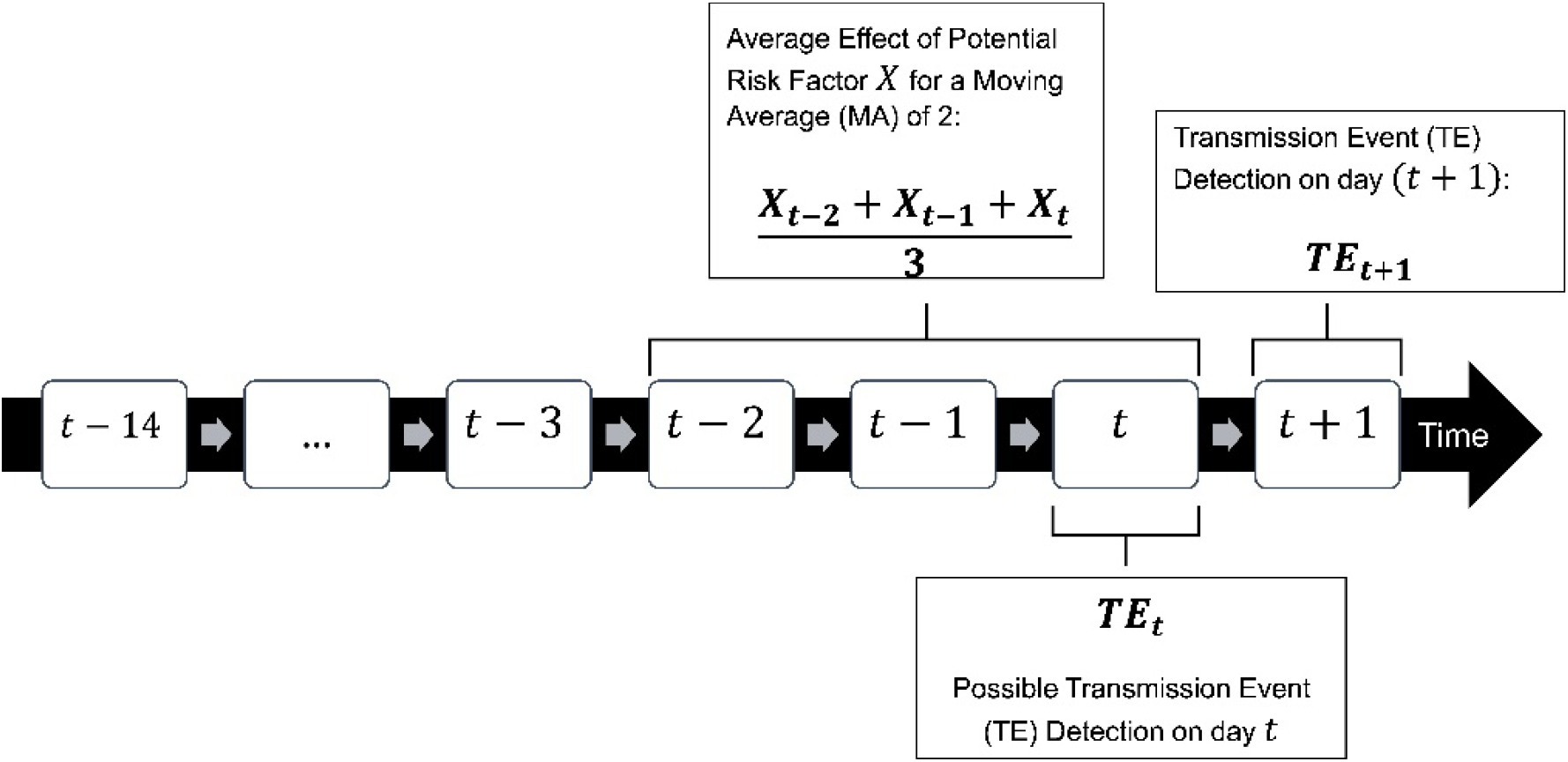
Illustration of the temporal relationships between the detection of transmission events and potential influencing variables X_i_. The variable TEs (transmission events) can only determine past transmissions, but not the exact time of the TE. In order to consider this measurement’s inaccuracy, the various independent variables X_i_ are also considered in different time periods with the aid of moving averages (MA).

## Results

### Demographics

434 out of 551 NICU admissions were included. 113 patients did not meet inclusion criteria. Four patients were excluded due to missing data. We observed 7940 patient days, of which 7037 had complete data for risk factor modeling. Among enrolled patients, 121 [27.9% (95% CI, 23.9%-32.3%)] were VLBW infants (median birth weight: 980 g, median gestational age: 28.7 weeks) with primary caesarian section as the most common delivery mode [52.1% (95% CI, 42.8%-61.2%)]. 313 [72.1% (95% CI, 67.7%-76.1%)] patients were non-VLBW infants (median birth weight: 2,590 g, median gestational age: 36.7 weeks). Hospital stay was longer for VLBW infants (median: 29 days) than for non-VLBW infants (median: 6 days) (*<u>eTable 3</u>*). Stratification of birth weight showed that lower birth weight was associated with longer hospital stays (*<u>eFigure 1</u>*).

### Colonization

225 of 434 of enrolled patients, i.e. 51.8 % (95% CI, 47.1%-56.5%), were colonized with at least one MDRO+; this corresponded to 28.3 (95% CI, 24.8-32.3) patients per 1,000 patient days. With 426 MDRO+ colonizations detected, the overall colonization rate was 53.7 (95% CI, 48.7-59.0) per 1,000 patient days, regardless of patient assignment.

Among MDRO+, *E. coli* was most frequent (n = 114) with a colonization rate of 14.4 (95% CI, 12.0%-17.2%) per 1,000 patient days (*Figure 3A*). Other MDRO+ with a colonization rate between 5 and 10 % were MSSA (n = 59), *K. oxytoca* (n = 54), *E. cloacae* (n = 53), *K. pneumoniae* (n = 49) and *K. oxytoca / Raoultella spp*. (n = 44) (*Figure 3A*). VLBW infants were most commonly colonized by two different MDRO+ species; non-VLBW infants were most commonly colonized by zero MDRO+ species. Five patients were colonized with up to six different MDRO+ (*Figure 3B*).

**Figure 3:**
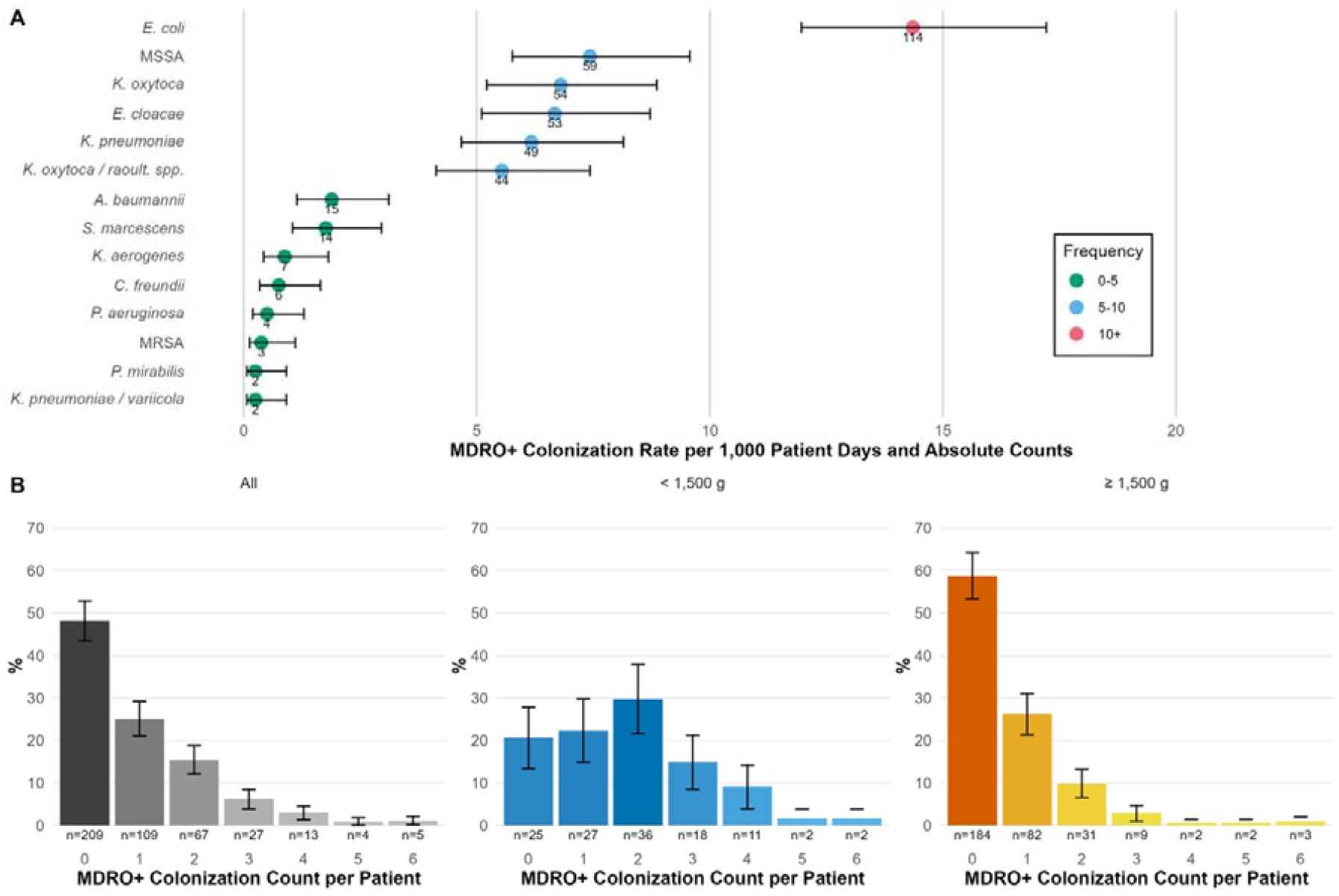
MDRO+ Colonization. **A)** Stratification of colonization rates per 1,000 patient days of MDRO+ by species with 95% CIs. Rates were attributed to frequency categories with color indicators. For every rate, the absolute count of respective MDRO+ isolates was stated. **B)** Distribution and counts of patients that experienced non-, single- or multiple MDRO+ colonizations with 95% CIs. Patient representation in groups: all (grey), birth weight < 1,500 g (blue), birth weight ≥ 1, 500 g (orange).

In non VLBW-infants MDRO+ were found significantly earlier in screening (median: 4 days) than in VLBW infants (median: 8 days) (*<u>eFigure 2A</u>*). Among patients free of MDRO+ colonization, the mean length of stay was 5.37 (95% CI, 4.79-5.95) days for non-VLBW infants and 9.96 (95% CI, 6.01-13.9) days for VLBW infants (*<u>eFigure 2B</u>*). MDRO+ species stratification showed no significant difference in days to first MDRO+ detection per patient (*<u>eFigure 3</u>*).

### Transmission Clusters

WGS identified 38 unique transmission clusters. *E. coli* clusters were most frequent (n = 11); *K. oxytoca* formed the largest cluster (19 patients); *S. marcescens* formed a single cluster (7 patients) (*<u>eTable 4</u>*). The TE distribution over time is shown in *<u>eFigure 4A</u>*. The overall transmission rate of MDRO+ was 17.8 (95% CI, 15.0-20.9) per 1,000 patient-days, with E. coli having the highest single rate of 6.9 (95% CI, 5.3-9.0) per 1,000 patient-days (*<u>eFigure 4B</u>*). WGS identified 157 bacterial isolates not contributing to TCs (i.e. “singletons”) (*<u>eTable 5</u>*). 179 isolates contributed to TCs (*<u>eTable 4</u>*). The phylogeny of *E. coli* isolates is shown in *<u>eFigure 5</u>*.

### Discriminatory Capabilities of WGS

We compared precision and validity of AFLP and WGS. 317 of the 348 isolates subjected to WGS were also analyzed by AFLP or *spa*-typing (after removal of copy strains). For example, the AFLP type “D” of E. coli corresponded to sequence type (ST) 141 in all cases, but could be divided into three different WGS clusters 201903_ST141, 201905_ST141 and 201908_ST141 (*Figure 4A, <u>eTable 6</u>*). Regardless of cluster composition, *E. coli* ST 141 was the predominant sequence type (*<u>eFigure 6</u>*), accounting for 42.4% (95% CI, 31.2%-54.4%) of cluster-contributing *E. coli* isolates, followed by *K. oxytoca* ST 176, accounting for 52.6% (95% CI, 37.3%-67.5%) of cluster contributing *K. oxytoca* isolates.

**Figure 4:**
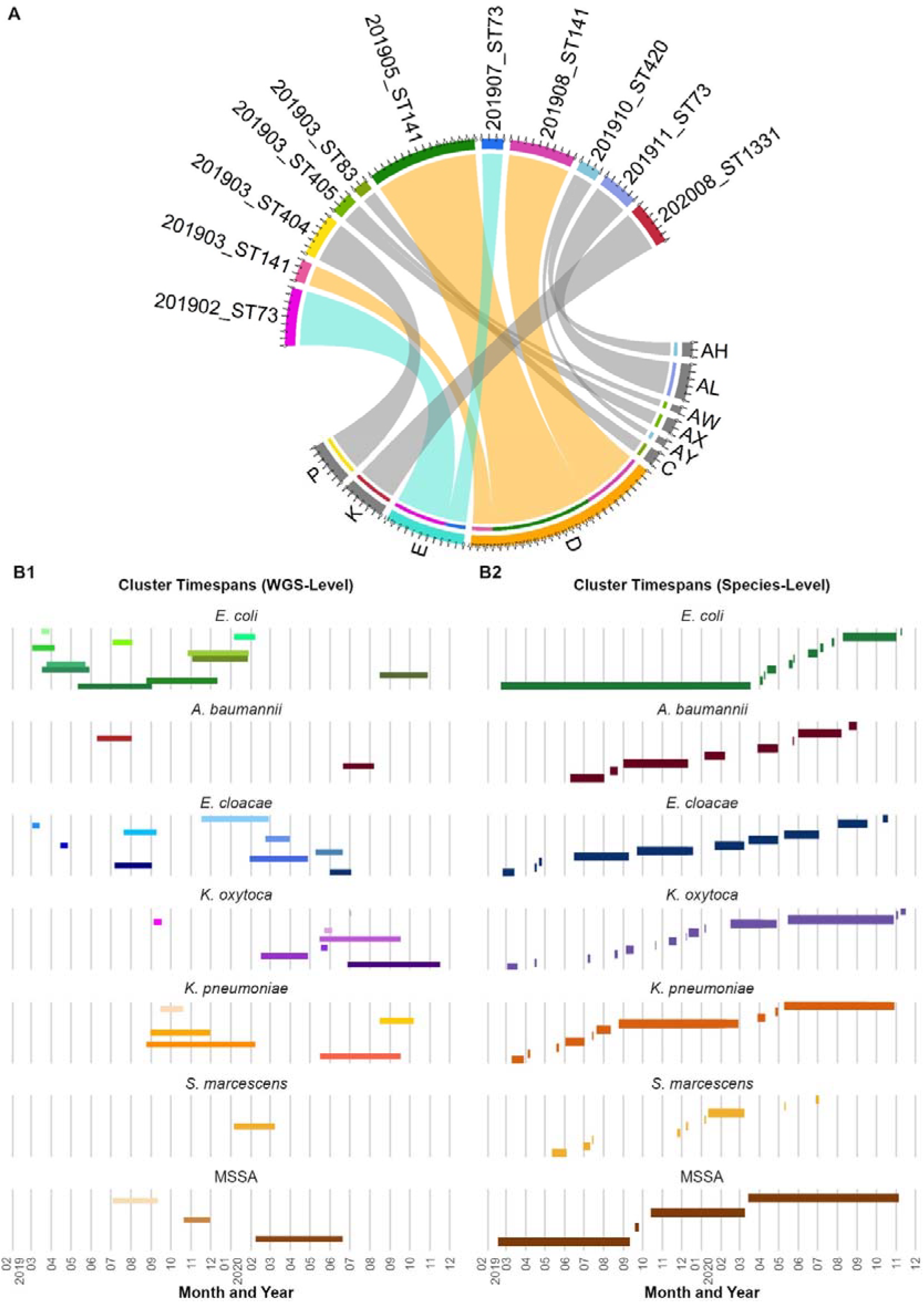
Variable Cluster Definitions. **A)** AFLP types (single- and dual-letters codes) were matched to their respective WGS clusters (year/month/underscore/ST/Number) in the chord diagram. Allocation of one AFLP type to various WGS-clusters was defined as a mismatch. For example: Perceivable mismatches for AFLP type “D” (orange) and “E” (turquoise), where one AFLP type was attributed to multiple WGS-clusters; perceivable mismatches for AFLP-type “AY” and “AH”, where both AFLP-types belonged to one WGS-cluster. **B1)** Cluster definition according to WGS: The combined timeframes of residing patients colonized with WGS-cluster-contributing isolates were represented over time and indicated by horizontal bars. The day of the index case detection defines the start of the timeline. For respective clusters, the discharge of the last patient colonized with a WGS cluster contributing isolate defined the end of the timeframe. Singletons (non-cluster contributing isolates) were excluded in this representation. Every species is represented in different shades of their respective base color, indicating different clusters (darker shades involve more cluster-contributing isolates). **B2)** Cluster definition according to species alone: Overlapping timeframes from both figures for respective MDRO+ indicated a gain in resolution in favor of WGS when compared to cluster determination by species alone, best exemplified by *E. coli*.

When comparing AFLP typing to WGS cluster identification, mismatch rates between the methods were 0.10, 0.09, and 0.06 for *K. oxytoca, E. cloacae*, and *E. coli* (*<u>eTable 7</u>*). Same species MDRO+ can cluster differently depending on the strain definition. For example, “*E. coli* Cluster 1” (lowest green bar in *Figure 4B2*) was defined as the temporal overlap of patients colonized with *E. coli* on the ward and formed a large cluster which can be further divided into distinct clusters as defined by WGS (*Figure 4B1, <u>eTable 8</u>*), leading to a resolution gain of 9 clusters. Increases in cluster resolution were observed for other species as well (e.g. “*K. oxytoca* cluster 12“; *<u>eTable 8</u>)*.

### MDRO+ Blood Stream Infections

Ten MDRO+ bloodstream infections (BSI) with *E. coli* [5], *K. pneumonia* [3] and *K. oxytoca* [2] were detected. Four patients had BSI derived from a TE (as revealed by WGS), of which three were detected with a same strain MDRO+ beforehand and admitted with a birth weight of < 1,000 g (*<u>eFigure 7</u>*). The BSI isolates belonged to WGS clusters 201907_ST73, 201911_ST73, 201908_ST219, 202006_ST966 (*<u>eTable 4</u>*). The overall rate of BSI was 1.3 (95% CI, 0.6-2.3) per 1,000 patient days and 0.5 (95% CI, 0.1-1.3) for infections resulting from a TE.

### Antibiotic usage

VLBW infants received selected antibiotics or their combinations proportionally and significantly more often than non-VLBW infants (*<u>eFigure 8A</u>*). However, they did not differ in the total number of administrations (*<u>eFigure 8B</u>*).

### Multivariate Model Analysis

Multivariate analysis combined patient, genomic, clinical and staffing data. The logistic regression model predicts the probability of becoming part of a TE (outcome variable) based on five predictor variables (*Figure 5A*). Of these, three predictor variables were significantly associated with the outcome variable (*Figure 5A, 5B*):

**Figure 5:**
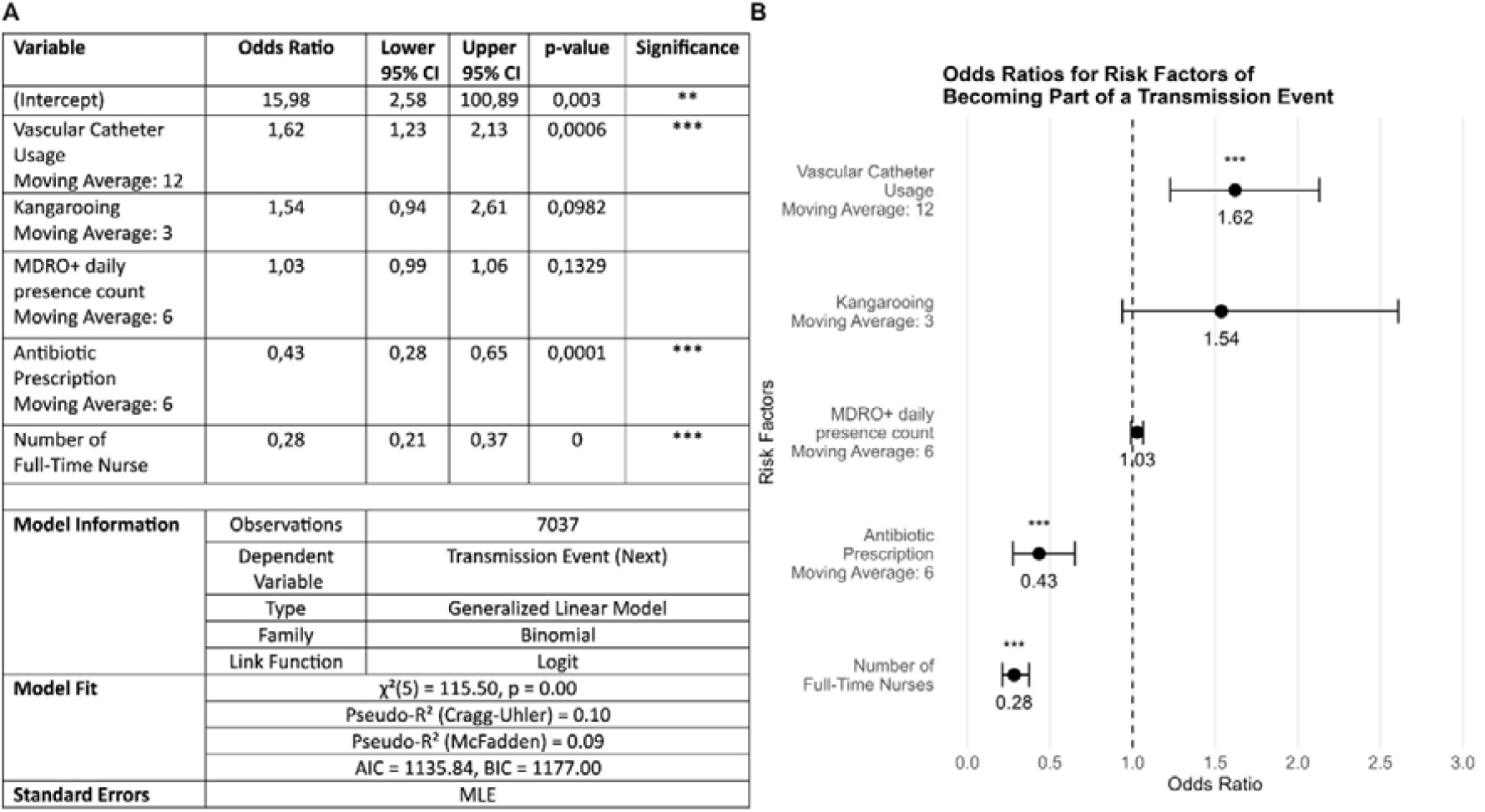
Multivariate Logistic Regression Model. **A)** Description of the proposed logistic regression model that aims to predict an infant’s probability of becoming part of a transmission cluster (outcome variable) based on five predictor variables with three significant associations out of five selected risk factors. The model type is a generalized linear model with a binomial family and logit link function. The model shows statistical significance (χ^2^(5) = 115.50, p < 0.001), indicating that the predictors, as a group, are generally associated with the outcome. **B)** Forest plot of odds ratios according to the logistic regression model. *: p-value < 0.05, **: p-value < 0.01, ***: p-value < 0.001

1. One-unit increase of the *Number of Full-Time Nurse*s decreases the odds by 72%.
2. One-unit increase of *Antibiotic Prescription*s (moving average 6) decreases the odds by 57%.
3. One-unit increase of *Vascular Catheter Usage* (moving average 12) increases the odds by 62%.

*Kangarooing* (moving average 3) and *MDRO+ daily presence count* (i.e. ‘colonization pressure’, moving average 6) were not significant. With a p-value < 0.001 for χ^2^(5), the final model outperformed the null model. However, overall fit was weak to moderate, as indicated by limited pseudo-R^2 (McFadden and Cragg-Uhlmann). Variance Inflation Factor (VIF) tests indicated no multicollinearity. A rainbow test for linearity indicated that not all logistic regression assumptions were met, limiting the results’ significance (*Figure 5A*).

## Discussion

This study identifies prospective WGS of routine screening isolates from newborns in intensive care as a powerful tool to detect and track transmission chains with MDRO+ at the highest precision. The transmission of – in part hospital-adapted – microorganisms between patients is commonly regarded as a failure of basic hygiene procedures, especially in high-risk individuals such as extremely premature infants. However, since very preterm infants typically depend on life-supporting measures immediately after birth, separation from the mother is the norm, i.e. microbiome development largely relies on input from staff skin and inanimate surfaces [23]. In this context, even the most advanced hygiene measures may not prevent transmissions of potentially harmful enterobacteria and firmicutes like staphylococci and streptococci [1]. Furthermore, early exposure to these species may be a prerequisite for long term tolerance and thus eventually beneficial [24]. On the other hand, phenotypic selection may promote the emergence of strains that harbor traits facilitating inter-individual spread, in-host expansion and invasion. This may lead to NICU outbreaks that are particularly feared, especially when bacteria carry resistance against first-line antibiotics [25]. It seems reasonable that real-time knowledge of the number of patients carrying a particular strain, the length of time the strain has been in a NICU, and the number of infants who develop clinical symptoms associated with strain carriage will enable the risk of TC to be calculated. Accordingly, accuracy and speed in bacterial typing should substantially improve accuracy of outbreak prediction [16]. However, due to the routine turnaround times for bacterial typing (including material shipment, deoxyribonucleic acid [DNA] preparation and typing), IPC measures are usually initiated immediately after the appearance of clusters of the same species (i.e. before typing results are available) and typing is often not initiated. Our surveillance findings align with national species-level data, emphasizing gram-negative bacteria (especially *E. coli, E. cloacae, K. oxytoca* and *K. pneumoniae*) as key screening targets and causes for nosocomial infections [5,26]. However, defining clusters only on the species level will inevitably mischaracterize TE and initiate unnecessary interventions such as ward closures, the effectiveness of which remains unclear and may compromise quality of care [27,28]. We found AFLP to detect TE in the majority of cases, similar to what has been reported for Fourier-transformed-infrared-spectroscopy-based typing [29]. However, both methods have limitations in cluster characterization and longitudinal surveillance, including considerable mismatch rates. Accordingly, we asked whether genomic cluster definition by WGS, the gold standard for bacterial strain discrimination [30,31], translates into a clinical advantage in a level III NICU. In fact, we found that WGS revealed AFLP-defined TC as pseudo-clusters. For example, *E. coli* formed multiple TC of ST 73 and ST 141, which, upon in-depth phylogenetic analysis with WGS, fell into distinct clusters, highlighting the advantage of WGS compared to methods that focus on housekeeping genes. In selected cases, WGS delineated the “microbial biography” from admission, to same strain colonization and occurrence in clinical samples (blood and tracheal aspirates). Importantly, swab-based screenings have limited sensitivity for TE detection since they are influenced by bacterial density on mucocutaneous surfaces. Moreover, the necessary minimal handling of the most fragile very preterm infants may impact sampling quality [32].

Any typing method relies on sufficient DNA quantity. Therefore, we enriched picked colonies by liquid culture. This may select for strains with growth advantages in culture (in the case of mixed strains of the same species), potentially impacting our results.

The precision in TC definition by WGS enabled us to identify factors in multivariate analysis affecting transmission. Most notably, the addition of only one full time nurse on the ward decreased the patients’ risk of becoming part of a TC. This seems by no means self-evident, since increasing personnel on a NICU may increase transmissions as well, e.g. due to more procedures, longer incubator opening times etc. Notably, staffing data included nurses declared as state-examined with high-qualification and did not account for nurses in training. Other training levels and professions should be included in further research. In order to investigate the potential of preventing or increasing TEs by increasing staffing levels, objective and highly granular quantification of hand hygiene measures is needed. In our hand hygiene compliance observations, we did not detect major changes in hand hygiene compliance over time. However, the data was not suitable for multivariate analysis. The presence of an indwelling catheters (arterial, venous) in patients increased the risk of becoming part of a TC. We hypothesize that vascular catheter presence may be linked to increased patient contact with staff (e.g. catheter care). Finally and to our surprise, prior administration of antibiotics reduced the risk of being part of a TC. Several factors may underlie this interrelation. The culture based screening method, preceding typing, may be impacted by antibiotics decreasing the bacterial density at the swabbed areas as shown for intestinal colonization [33,34]. These considerations are in line with the fact that VLBW patients received more antibiotics and were detected later for first MDRO+ colonization compared to non-VLBW patients. Given the evidence highlighting the potential short- and long-term effects of antibiotics, e.g. an increased risk for bronchopulmonary dysplasia [35], necrotizing enterocolitis or invasive fungal infections [36], atopy, asthma and even mortality, this finding needs exploration in independent studies.

In contrast to the outlined factors, number of patients colonized by MDRO+ on the NICU (‘colonization pressure’), kangarooing and the *INPULS* score [37] were not significantly associated with TC. In the final model, the necessary condition of linearity cannot be met, which limits the validity of the model. This exemplifies the complexity of identifying practically useful risk factors with different temporal dynamics. Non-linear alternatives such as fractional polynomials [38–40], splines [41] or non-linear distributed lag models [42] are not feasible in clinical practice due to the lack of interpretability or they are significantly more complex and less robust due to the additional assumptions that need to be made.

In summary, we identify WGS, as part of the colonization screening, to be of great value in systematically resolving MDRO+ transmission chains with the highest precision. Prospective integration of granular patient and staffing data with multivariate model analysis builds the foundation for rational, data-driven and actionable conceptualizations of IPC-measures in the future.

## Data Availability

Due to confidentiality restrictions, the data supporting this study are not publicly available. However, they are available from the corresponding author upon reasonable request.

## Author Contributions

Tim Götting and Nico Mutters were the initial investigators, published the initial study protocol and transferred data and responsibility to Timmy Nguyen and Fabian Bürkin after leaving the Institute for Infection Prevention and Control at the Medical Center – University of Freiburg, Germany.

### Concept and design

Tim Götting, Nico Mutters, Timmy Nguyen, Philipp Henneke, Sandra Reuter

### Acquisition of data

Tim Götting, Timmy Nguyen, Nico Mutters

### Analysis and interpretation of data

Timmy Nguyen, Fabian Bürkin, Stefany Ayala-Montaño, Tim Götting, Philipp Henneke, Sandra Reuter

### Provision of microbiological isolates

Christian Schneider

### Genomic and molecular analysis

Stefany Ayala-Montaño, Iván Acevedo Monterrosa, Sandra Reuter, Daniel Jonas

### Multivariate statistical analysis

Fabian Bürkin, Martin Wolkewitz, Tjibbe Donker

### Drafting of the manuscript

Timmy Nguyen, Philipp Henneke, Fabian Bürkin

### Critical review of the manuscript for important intellectual content

Iván Acevedo Monterrosa, Daniel Jonas, Daniel Klotz, Hans Fuchs, Martin Kuntz, Christian Schneider, Martin Wolkewitz, Tjibbe Donker, Sandra Reuter, Tim Götting

### Supervision

Philipp Henneke, Tim Götting

## Acknowledgements

We sincerely thank Leonardo Duarte dos Santos from Group Reuter and Sabine Weber, Marion Buck, Sophie Sträb, Doris Scheibert and Eva Zimmermann from Group Jonas and Arvid Dürkop as NICU head nurse for their invaluable contributions and support.

## Ethics and dissemination

The study was approved by the Ethics Committee, Medical Center – University of Freiburg, Germany, on 28 August 2018 (registration number 287/18).

## Registration

The study was registered in the German Clinical Trials Register (DRKS) under the registration ID DRKS00017733.

## Abbreviations

*Abbreviation Meaning*

NICU: Neonatal intensive care unit
TE: Transmission event
MDR: Multi drug resistant / multi drug resistance
NI: Nosocomial infection
IPC: Infection prevention control
VLBW: Very low birth weight
RKI: Robert-Koch-Institute
MRSA: Methicillin resistant *Staphylococcus aureus*
MDRO+: Multidrug resistant organism plus
AFLP: Amplified fragment length polymorphism
WGS: Whole genome sequencing
TC: Transmission cluster
G-BA: Gemeinsamer Bundesausschuss
MSSA: Methicillin susceptible *Staphylococcus aureus*
CI: Confidence interval
ST: Sequence Type
BSI: Blood stream infection
VIF: Variance inflation factor
DNA: Deoxyribonucleic acid

## Supplement

PROtectNEO_Preprint_SUPPLEMENT.pdf

PRJEB81699_Accessions.xlsx

