## Supplement for "Prospective whole-genome sequencing uncovers factors influencing bacterial transmission in neonates (PROtect NEO)"

#### Table of Contents

##### Methods


eMethod 1: Routine Cultural Diagnostics

eMethod 2: Collection and Preparation of Isolates

eMethod 3: Collection of Nurse Staffing Data

eMethod 4: Data Management

eMethod 5: Genetic Typing of Bacterial Strains

eMethod 6: Statistical Analysis


##### Abbreviations

##### References

##### Figures


eFigure 1: Length of Stay of Patient Population

eFigure 2: MDRO+ Colonization Dynamics on Patient Level

eFigure 3: MDRO+ Colonization Dynamics on Species Level

eFigure 4: MDRO+ Transmissions

eFigure 5: Phylogeny of *E.coli* Isolates

eFigure 6: Cluster Stratification by Sequence Type

eFigure 7: Alluvial Diagram of observed MDRO+ Blood Stream Infections

eFigure 8: Antibiotic Administration by Birth Weight Groups

#### Tables

eTable 1: Case-Report-Form

eTable 2: Candidate Set for Potential Confounders $X_{(i, t)}$

eTable 3: Demographic Data of Study Population

eTable 4: WGS-Cluster-Contributing MDRO+

eTable 5: WGS-non-Cluster-Contributing MDRO+ ("singletons")

eTable 6: AFLP-Mismatches for *E. coli*

eTable 7: Cumulative AFLP-Mismatch Rates per MDRO+

eTable 8: WGS Enabled Net Gain in Resolution

### Methods

#### eMethod 1: Routine Cultural Diagnostics

Screening swabs (eSwab, Copan, Brescia, Italy) and clinical samples were inoculated on both selective and non-selective media including the following agar plates: Columbia Blood, MacConkey, Hematin Chocolate Agar, chromID® (Biomérieux, Nuertingen, Germany) Extended spectrum beta lactamase (ESBL) and Methicillin resistant *Staphylococcus aureus* (MRSA) selective. Media were incubated under aerobic conditions for 48 hours at 36°C and with 5% $CO_2$. If growth on plates was detected, matrix-assisted-laser-desorption-ionization-time-of-flight (MALDI-TOF) mass spectrometry (Bruker Daltonics, Bremen, Germany) was employed to identify bacterial species. Susceptibility testing was performed using VITEK2® (Biomérieux, Nuertingen, Germany) and/or minimum inhibitory concentration (MIC) test strips (Liofilchem, Piane Romano, Italy) respectively. The results were interpreted according to European Committee on Antimicrobial Susceptibility Testing (EUCAST) clinical breakpoints. Presence of resistance genes was confirmed by nucleic acid amplification tests for carbapenem-resistant gram-negative bacteria and MRSA.

#### eMethod 2: Collection and Preparation of Isolates

If multidrug resistant organism plus (MDRO+) species were found, bacterial isolates were transferred to the typing laboratory. The study included the following bacteria in our genetic and statistical analysis and are referred to as “MDRO+”, specifically: *A. baumanii, C. freundii, E. coli, E. cloacae, K. oxytoca, K. pneumonia, P. mirabilis, S. marcescens, P. aeruginosa* and *S. aureus* (methicillin sensitive *Staphylococcus aureus* [MSSA], MRSA) on the basis of the initially published study protocol [1].

#### eMethod 3: Collection of Nurse Staffing Data

Procedural data contained anonymized daily and shift-specific metrics on bed occupancies, nurse staffing levels and application of minimum staffing criteria. The patients’ birth weight, maturity level and complexity of intensive care treatment or intensive care monitoring guided definitions of ‘overstaffing’ and ‘understaffing’ at shift level, which are based on algorithms predefined by the Federal Joint Committee, a national regulatory authority which issues mandatory directives on above mentioned criteria (G-BA, German: “Gemeinsamer Bundesausschuss”) [2].

#### eMethod 4: Data Management

Data was collected with Microsoft *Access*, further analyzed with Microsoft *Excel* as well as the statistical Software *R* (Version 4.4.1). Patient data pseudonymization via hash-functions. We collected clinical data from patient charts or electronic systems (laboratory information system, clinical information system) including patient- and ward-specific data, potential risk factors and confounders as well as procedural and operational data. Information on the case-report-form is provided (*eTable 1*). Outliers and influential values were not corrected or excluded, as extreme situations were considered particularly interesting for the analysis due to everyday hospital hygiene practice and their clinical relevance.

#### eMethod 5: Genetic Typing of Bacterial Strains

Amplified fragment length polymorphism (AFLP) typing for gram-negative bacteria, *spa*-typing for *S. aureus* (MSSA, MRSA) and Whole genome sequencing (WGS) were performed for typing of MDRO+.

*AFLP*

AFLP typing from pure bacterial cultures was performed with the Genetic Analyzer abi3500. The comparison of the resulting fragment patterns was software-assisted (abi3500 analysis software *GeneMapper*™ [Thermo Fisher Scientific], *BioNumerics* [Applied Maths NV]). The assignation of fragment differences to genotypes was performed as described [3,4].

*WGS*

Deoxyribonucleic acid (DNA) extraction was performed with a High Pure polymerase chain reaction (PCR) Template (Roche) following standard protocol. Sequencing was carried out using the Illumina MiSeq Nextera DNA Flex Library pre-preparation and V2 300 cycle PE kit according to manufacturer's instructions. Bioinformatics analysis was carried out using smalt [5], samtools v0.1.19 [6] and GATK (mapping) [7], SPAdes v3.13.1 [8] with kmer sizes 21, 33, 55, 77, 99, 109, and 123 with filtering to only include contigs with a minimum of 500bp (assembly), kraken v1.1.1 [9] (species identification), mlst v2.10 [10] (multilocus sequence typing [MLST] identification). Quality control parameters were a minimum of 30x coverage, appropriate length of

the sequence, number of contigs <500, N50 <100,000bp, correct species and MLST identification. Phylogenetic reconstruction was based on core genome alignment, single nucleotide polymorphism (SNP) identification (snp-sites v2.5.1, snp-dists v0.6), followed by estimation using RAxML v8.2.12. iTOL was used for phylogenetic visualization. Sequencing data has been deposited in the ENA project PRJEB81699 and individual accession identifiers can be found in the supplemental material.
If bacterial isolates are indistinguishable from one another via WGS, based on respective cut-off values, we interpret them as resulting from transmission events. The first chronological appearance of an isolate in a cluster was regarded as the index case. The minimum number of indistinguishable bacterial isolates required to form a cluster is two.

#### eMethod 6: Statistical Analysis

Following Barnett & Graves [11] and Breslow et al. [12], temporal dynamics of the multivariate risk factor analysis for transmission events (TE) was mapped using a logistic regression model, which allows a differentiated view of relevant time periods before a TE could be detected. We also analyzed mixed models. However, the analysis was conducted based on logistic models since random parts (patients, time and both) were not significant.

*Dependent Variable $Y_t$: Transmission Events*

The dependent variable $Y_{t+1}$ is a binary representation of the TE at time *t+1* ($TE_{(t+1)}$) at the individual patient level with a value of 1 for a day with positive MDRO+ screening and 0 on days with negative or no MDRO+ screening. The postponement of Y in the form of *t+1* instead of t has practical reasons: If a screening is positive on day *t+1*, with the incubation period the transmission took place on day *t* at the latest, especially since a large proportion of the screenings took place in the morning.

*Independent Variables $X_{(i, t)}$*

The candidate set for potential confounders $X_{(i, t)}$ with *i in {1, …, 18}* in the model included 18 variables (*eTable 2*). The variables $X_{(i, t)}$ were also considered as equally weighted moving averages (MA) over the time periods *[t; t], [t; t-1], [t; t-2], ..., [t; t-14]*, so that a total of 15 temporally nuanced variants of each variable were available for selection.

For *d* in *{0,1, ..., 14},* the respective moving average was defined as:

$$\text{MA: d} := \sum_{j=t}^{t-d} \frac{X_{(i,j)}}{d+1}.$$

In our model e.g. "MA: 2" represents the unweighted average over the two days *t*, the previous day *t-1* and *t-2* (*Figure 2*).
An optimal model was chosen using minimum Akaike's information criterion (AIC). Predictors were selected via stepwise forward-backward selection. Starting from a null model, all univariate models were tested, and the best combination of a confounder and moving average (yielding lowest AIC) was selected. To ensure interpretability, we excluded all alternative moving averages of that predictor among all available variables. If several variants of predictors measured the same target variable (e.g. staff occupancies), only one variable was retained. After adding a new predictor, each included variable was re-evaluated using a 'leave-one-out' approach to seek a lower AIC score alternatives. This process was repeated until no further improvement was possible after including at least two additional variables. The final model, with the lowest AIC, was then tested for multicollinearity (using variance inflation factor [VIF]) and linearity (using a rainbow test [13,14]) to meet logistic regression conditions.

*Conditions for logistic regression*

One of several necessary conditions for logistic regression is that of linearity. For our model, this condition is checked by performing the rainbow test. In order to restore the condition of linearity when the rainbow test is significant, transformations such as Box-Cox [15] or Yeo-Johnson [16] would be necessary from a technical perspective. However, back transformations of the variables are not valid and the cumbersome interpretations of the transformations prevent a meaningful application in clinical practice. For this reason, even if the rainbow test is significant, no further measures are taken in favor of the interpretability of the results, even if this limits the validity of the model itself.

#### Abbreviations

| *Abbreviation* | *Meaning* |
|---|---|
| ESBL | Extended spectrum betalactamase |
| MRSA | Methicillin resistant *Staphylococcus aureus* |
| MALDI-TOF | Matrix-assisted-laser desorption-ionization time-of-flight |
| MIC | Minimum inhibitory concentration |
| EUCAST | European Committee on Antimicrobial Susceptibility Testing |
| MDRO+ | Multidrug resistant organism plus |
| MSSA | Methicillin susceptible *Staphylococcus aureus* |
| G-BA | Gemeinsamer Bundesausschuss |
| AFLP | Amplified fragment length polymorphism |
| WGS | Whole genome sequencing |
| DNA | Deoxyribonucleic acid |
| PCR | Polymerase chain reaction |
| MLST | Multilocus sequence typing |
| SNP | Single-nucleotide polymorphism |
| TE | Transmission Event |
| MA | Moving averages |
| AIC | Akaike’s information criterion |
| VIF | Variance inflation factor |

#### Figures

##### eFigure 1: Length of Stay of Patient Population

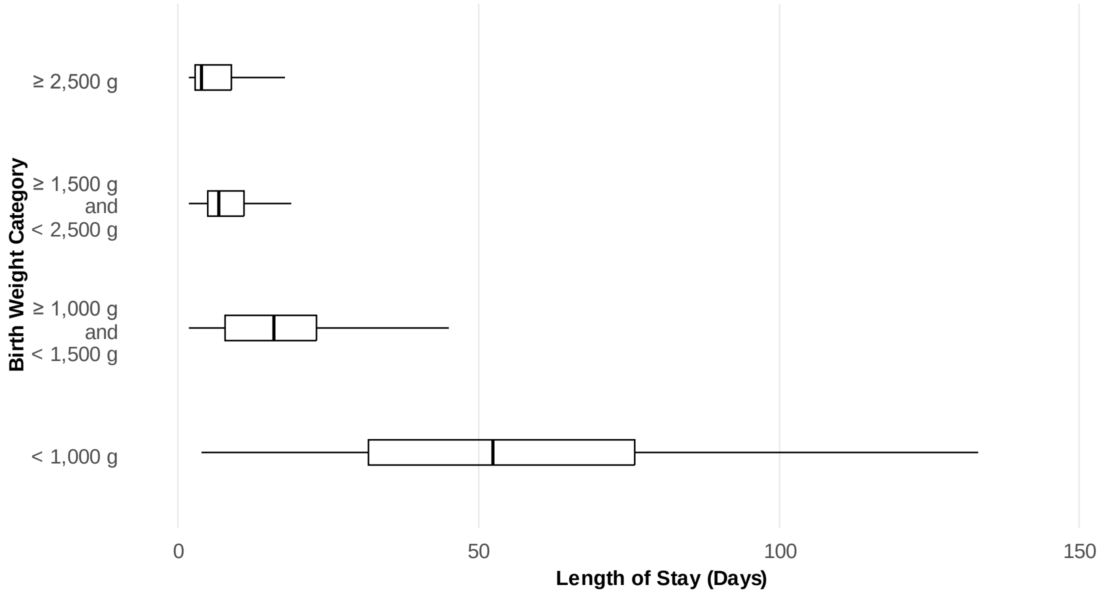


Patient length of stay in days, stratified by birth weight categories.

### eFigure 2: MDRO+ Colonization Dynamics on Patient Level

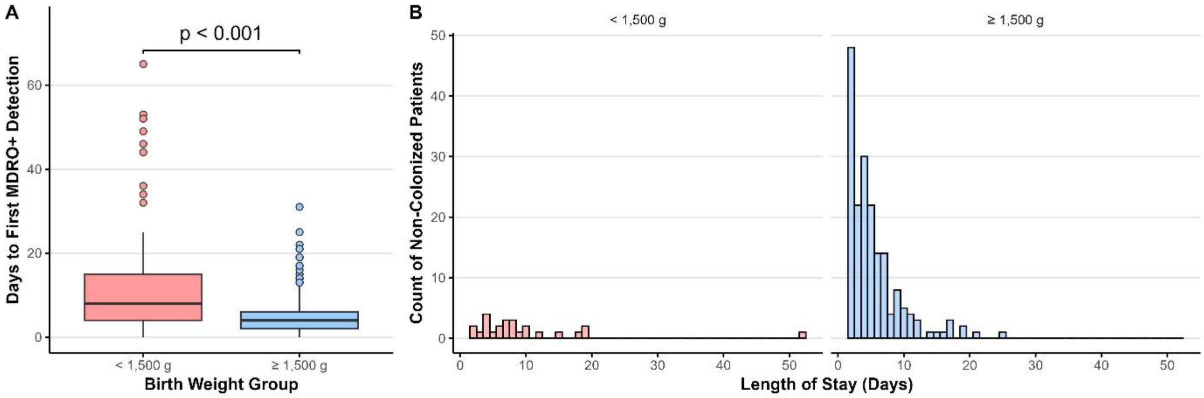


**A)** Number of days from admission to first MDRO+ detection (regardless of species) by birth weight groups. Patients with birth weight ≥ 1,500 g were colonized with their first MDRO+ significantly earlier (Wilcoxon Rank-Sum Test, $p < 0.001$). **B)** Distribution of length of stay of patients that were not colonized by MDRO+ and stratification by birth weight.

#### eFigure 3: MDRO+ Colonization Dynamics on Species Level

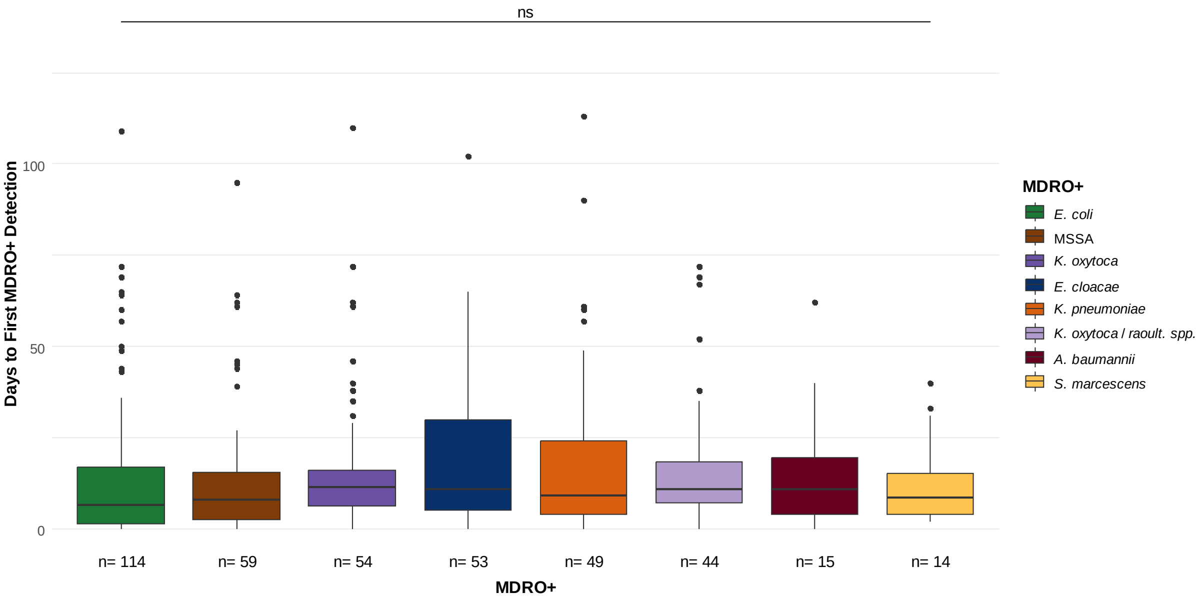


Number of days from admission to first MDRO+ detection (stratified by MDRO+ species). The comparison of MDRO+ showed no statistically significant differences across all combinations of MDRO+ for the time to first detection (Kruskal-Wallis chi-squared = 12.863, df = 7, p = 0.076).

### eFigure 4: MDRO+ Transmissions

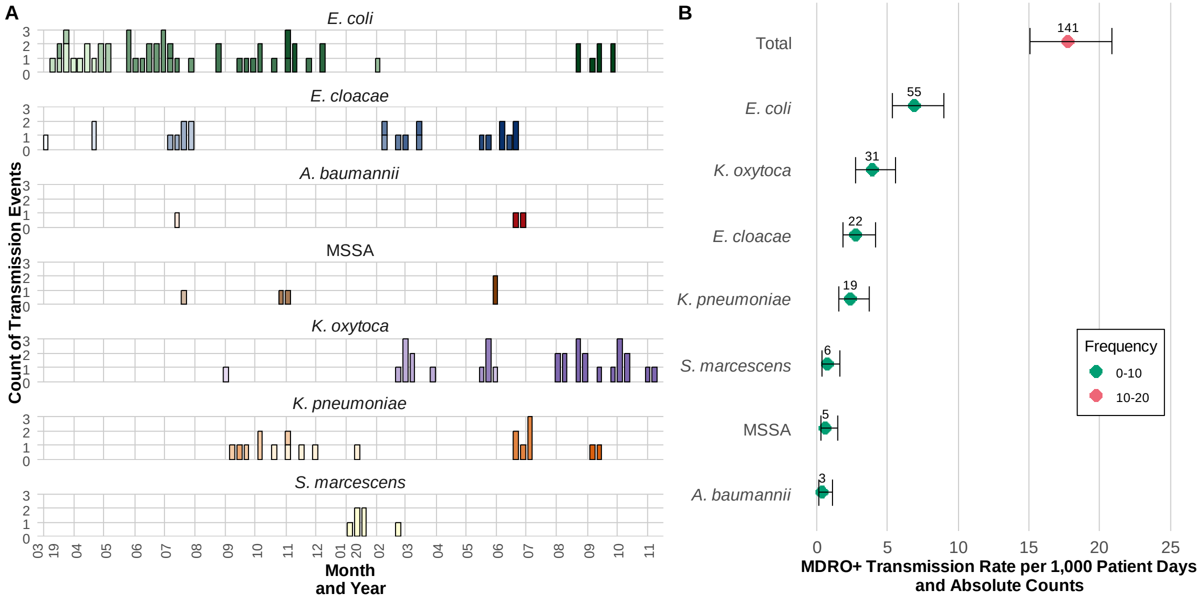


**A)** Occurrence of transmission events over time, stratified by MDRO+ species. Every MDRO+ species is displayed in their respective base colors. Shades of colors indicate the attribution to separate cluster as defined by WGS. Cluster-index-cases were excluded in this representation. **B)** Transmission rate per 1,000 patient days with 95% CIs and category attribution. Stratification by MDRO+ species and in total with respective absolute counts.

#### eFigure 5: Phylogeny of *E.coli* Isolates

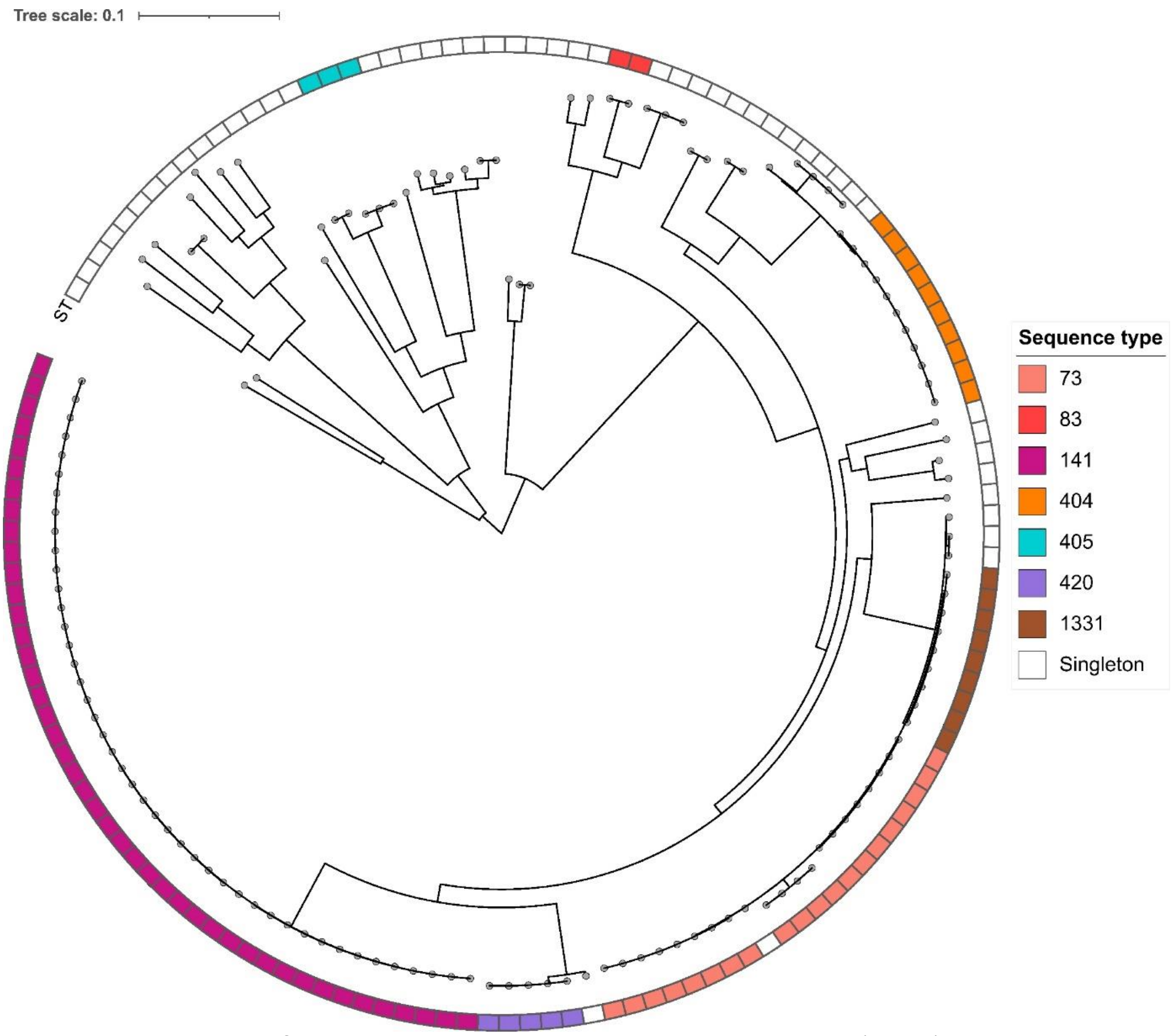


*E.coli*. was the dominant MDRO+ species in our study, comprising several cluster contributing (colored) and non-cluster-contributing, i.e. "singletons" (white) isolates with respective sequence types (ST). ST141 was the dominant sequence type (n=28) that could further be differentiated into separate WGS-clusters.

### eFigure 6: Cluster Stratification by Sequence Type

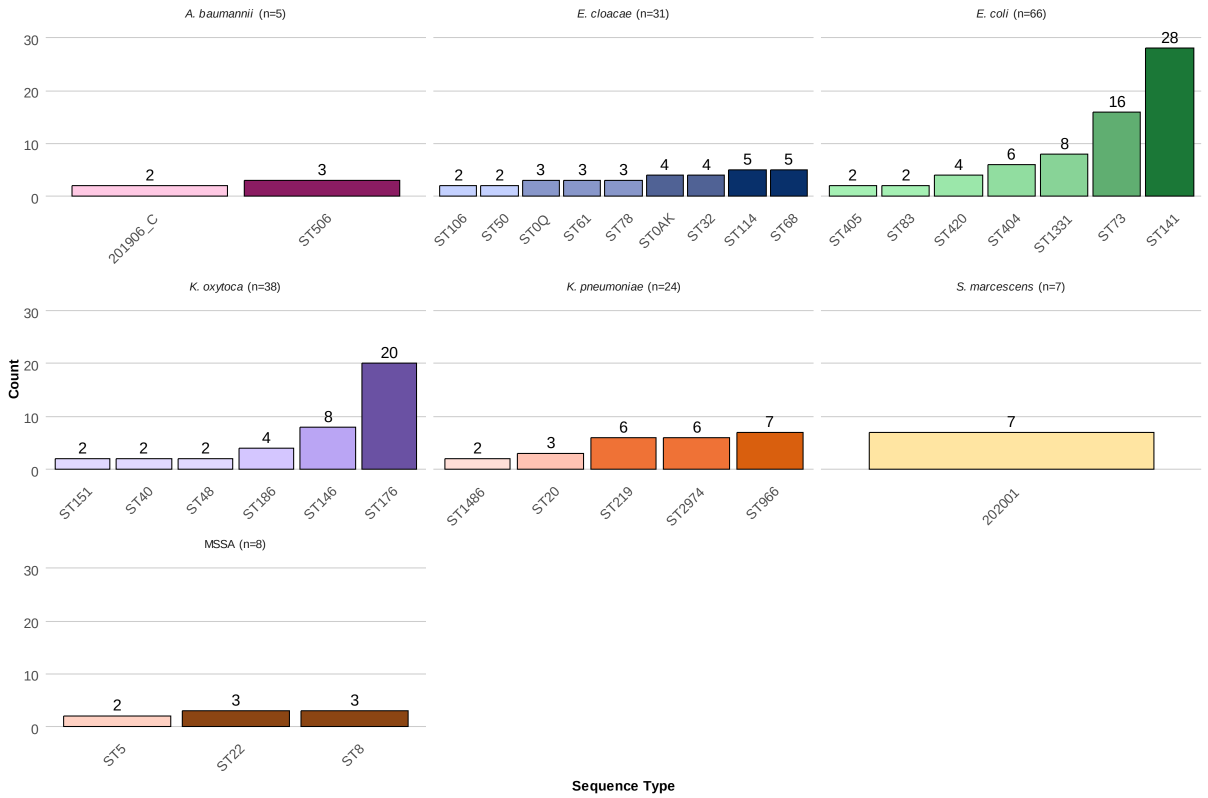


Stratification of cluster contributing MDRO+ isolates on sequence type level. The three most dominant species in our study were (descending): *E. coli*, *Klebsiella spp*. and *E. cloacae*.

#### eFigure 7: Alluvial Diagram of observed MDRO+ Blood Stream Infections

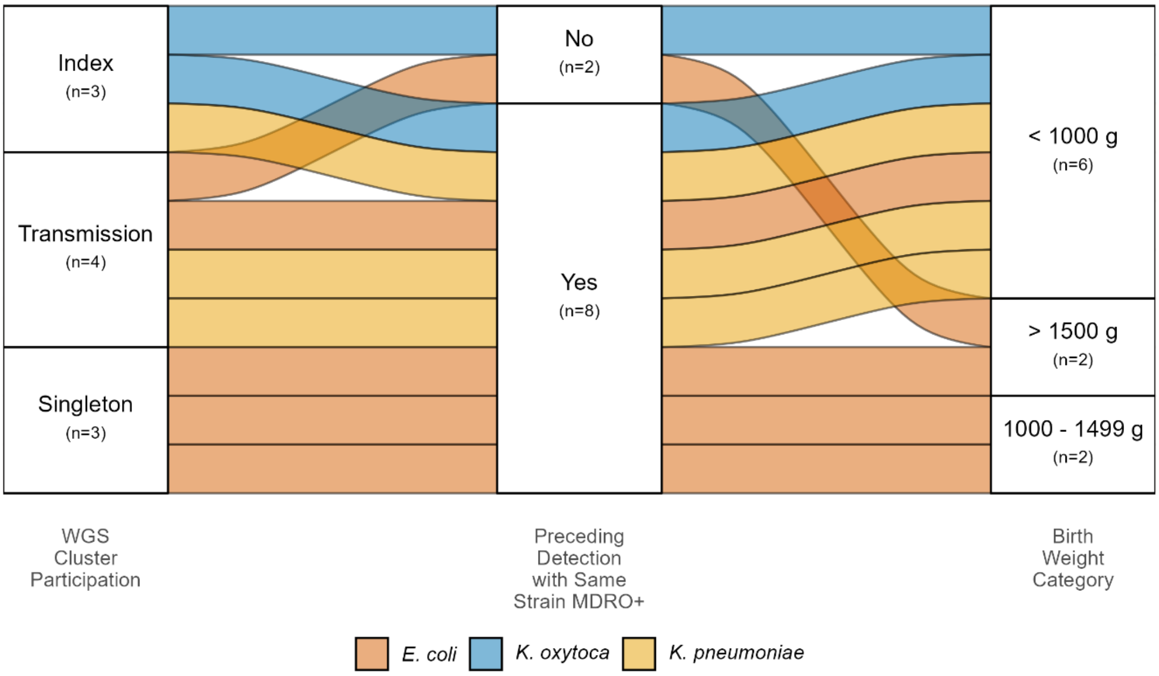


Every combination of three categories, indicated by connecting lines, resembles one patient. Seven BSI could be attributed to a TC as defined by WGS (Index, Transmission), of which 4 derived from a TE. Of these, three were preceded with the detection of same strain MDRO+ in materials other than blood culture samples.

#### eFigure 8: Antibiotic Administration by Birth Weight Groups

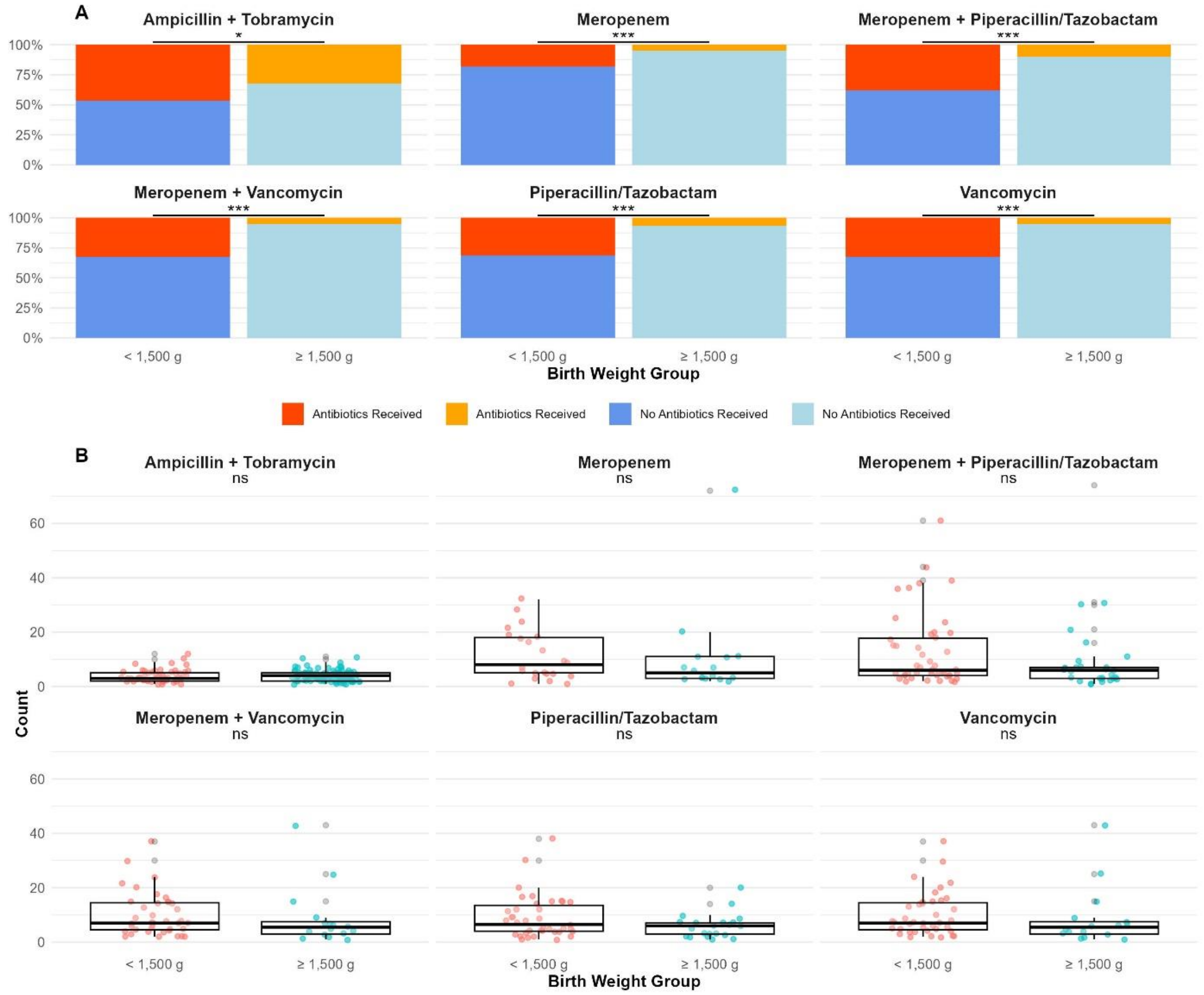


**A)** Proportion of patients treated with antibiotics or their combinations vs. patients that did not receive any antibiotics (stratified by birth weight groups). Proportionally, patients with a birth weight < 1,500 g receive the selected antibiotics or their combinations more often. Statistical significance according to Chi-Squared Test (* $p < 0.05$, ** $p < 0.01$, *** $p < 0.001$). **B)** Counts of antibiotics administered per patient, stratified by birth weight group. No statistical differences between groups and administered antibiotics or their combinations according to Wilcoxon Rank-Sum Test (ns = not significant).

### Tables

#### eTable 1: Case-Report-Form

| ***CRF*** | |
|---|---|
| ID | Vancomycin |
| PIZ (Pseudonym) | Tazobactam |
| Patient Number | Cotrim |
| Patient Name (Pseudonym) | Meropenem |
| Patient First Name (Pseudonym) | Cefuroxim |
| Date of Birth | Ampicillin |
| Sex | Tobramycin |
| Birth Weight | Erythromycin |
| Mode of Delivery | Amphotericin B |
| Multiple Birth | Rifampicin |
| Observation Date | Cefotaxim |
| Admission | Levofloxacin |
| Discharge | Teicoplanin |
| Type of Discharge | Linezolid |
| Room | Ceftazidim |
| Bed Location | Material |
| Incubator Number | Swab Site |
| Invasive Breathing | MDRO+ |
| CPAP | Case Number (Microbiology) |
| CVC | Sample Number (Microbiology) |
| PVC | Date of Report (Microbiology) |
| UVC | Date of Sample Entry (Microbiology) |
| Arterial Line | Date of Report (AFLP) |
| Breast Milk | Internal Isolate Code |
| Feeding Tube | Resistence Category (KRINKO) |
| Bladder Catheter | Report Number (AFLP) |
| EVD | AFLP-Type |
| Kangarooing | Sequence Type (WGS) |
| INPULS Score | Cluster Definition (WGS) |

Column headers of the Case-Report-Form.

#### eTable 2: Candidate Set for Potential Confounders $X_{(i,\,t)}$

| **Individual Variables – Per Patient** | | |
|---|---|---|
| ***Variable*** | ***Type*** | ***Explanation*** |
| Sex | Binary | Sex of patient |
| Delivery | Categorical | Type of birth (C-section emergency, C-section primary, C-section secondary, vaginal) |
| Birth Weight | Continuous | Birth weight of patient in grams |
| INPULS* | Continuous | Based on categorical severity score INPULS® in intensive care units (treated as a continuous variable in order to be able to map different temporal dynamics via moving averages) |
| Breast Milk | Binary | 1 if the patient received breast milk, 0 if not |
| Feeding Tube | Binary | 1 if the patient had a feeding tube in place, 0 if not |
| Kangarooing | Binary | 1 if the patient experienced kangarooing, 0 if not |
| Antibiotics | Continuous | Number of different antibiotics administered, regardless of dose and frequency (Ampicillin, Tobramycin, Piperacillin/Tazobactam, Meropenem, Vancomycin, Cefuroxim, Erythromycin, Rifampicin, Cefotaxim, Levofloxacin, Teicoplanin, Linezolid, Ceftazidin, Cotrim, Amphotericin B) |
| Catheter | Continuous | Number of different catheters (venous [peripheral, central] and arterial) |
| Invasive Ventilation | Binary | 1 if the patient was ventilated with a breathing tube, 0 if not |
| **Environmental Variables – Per Ward** | | |
| ***Variable*** | ***Type*** | ***Explanation*** |
| MDRO+ | Continuous | Total number of MDRO+ present per day |
| Bed Occupancy | Continuous | Number of patients, averaged over all three shifts |
| Nurse Patient Ratio | Continuous | Nurse patient ratio, averaged over all three shifts |
| Staff: FTE | Continuous | Nursing staff totalled across all three shifts in full-time equivalents (FTE) |
| Staff: Deviance | Continuous | Deviation between the nursing staff required by the G-BA and the staff actually deployed in full-time equivalents, averaged over all three shifts |
| Staff: Understaffed | Continuous | Number of missing full-time nursing staff compared to the requirement determined by G-BA, totalled across all three shifts |
| Staff: Overstaffed | Continuous | Number of additional full-time nursing staff compared to the requirement determined by the G-BA, totalled across all three shifts |

Differentiation of potential confounders for multivariate analysis in individual (patient specific) and environmental variables (ward specific). Potential confounders were also considered as equally weighted moving averages over the time (up to 14 days in the past).

*INPULS Score: Developed by University Hospital Heidelberg, the score is a standardized tool to quantify patients' nursing care needs by assessing a range of clinical and functional parameters, ranging from categories 1 for the lowest care needs to category 6 for the highest.

#### eTable 3: Demographic Data of Study Population

| | Overall | | Combined: < 1,500 g | | Combined: ≥ 1,500 g | |
|---|---|---|---|---|---|---|
| | N = 434 | | N = 121 | | N = 313 | |
| *Category* | *Count (%)* | *95% CI* | *Count (%)* | *95% CI* | *Count (%)* | *95% CI* |
| Gender | | | | | | |
| Female | 192 (44.24) | 39.52 - 49.06 | 52 (42.98) | 34.11 - 52.29 | 140 (44.73) | 39.16 - 50.43 |
| Male | 242 (55.76 ) | 50.94 - 60.48 | 69 (57.02) | 47.71 - 65.89 | 173 (55.27) | 49.57 - 60.84 |
| Birth Weight Category | | | | | | |
| < 1000 g | 64 (14.75) | 11.62 - 18.52 | 64 (52.89) | 43.64 - 61.96 | 0 (0) | NA |
| ≥ 1000 g, < 1500 g | 57 (13.13) | 10.17 - 16.76 | 57 (47.11) | 38.04 - 56.36 | 0 (0) | NA |
| ≥ 1500 g, < 2500 g | 146 (33.64) | 29.25 - 38.33 | 0 (0) | 0.00 - 3.8 | 146 (46.65) | 41.04 - 52.34 |
| ≥ 2500 g | 167 (38.48) | 33.91 - 43.26 | 0 (0) | 0.00 - 3.8 | 167 (53.35) | 47.66 - 58.96 |
| Delivery Mode | | | | | | |
| NA | 1 (0.23) | 0.01 - 1.48 | 1 (0.83) | 0.04 - 5.19 | 0 (0) | NA |
| C-section: Emergency | 35 (8.06) | 5.76 - 11.14 | 13 (10.74) | 6.07 - 18.00 | 22 (7.03) | 4.56 - 10.60 |
| C-section: Primary | 169 (38.94) | 34.36 - 43.72 | 63 (52.07) | 42.84 - 61.16 | 106 (33.87 ) | 28.69 - 39.44 |
| C-section: Secondary | 91 (20.97) | 17.29 - 25.17 | 30 (24.79) | 17.60 - 33.62 | 61 (19.49) | 15.34 - 24.41 |
| Vaginal | 138 (31.80) | 27.48 - 36.44 | 14 (11.57) | 6.70 - 18.97 | 124 (39.62) | 34.20 - 45.29 |
| Multiple Birth Mode | | | | | | |
| Twins | 79 (18.20) | 14.75 - 22.23 | 27 (22.31) | 15.46 - 30.97 | 52 (16.61) | 12.76 - 21.31 |
| Triplets | 7 (1.61) | 0.71 - 3.44 | 3 (2.48) | 0.64 - 7.62 | 4 (1.28) | 0.41 - 3.46 |
| Quadriplets | 3 (0.69) | 0.18 - 2.18 | 3 (2.48) | 0.64 - 7.62 | 0 (0) | NA |
| Single | 345 (79.49) | 75.32 - 83.13 | 88 (72.73) | 63.75 - 80.24 | 257 (82.11) | 77.31 - 86.10 |
| *Category* | Median (Mean) | IQR (min - max) | Median (Mean) | IQR (min - max) | Median (Mean) | IQR (min - max) |
| Birth Weight (grams) | 2,165 (2,215) | 1,549 (405 - 4,870) | 980 (1,019) | 555 (405 - 1,490) | 2,590 (2,678) | 1,230 (1,520 - 4,870) |
| Gestational age (weeks) | 34.6 (34.27) | 6.9 (22.4 - 42.4) | 28.7 (28.6) | 4.6 (22.4 - 34.9) | 36.7 (36.4) | 5.14 (30.0 - 42.4) |
| Length of Stay (days) | 8.0 (17.8) | 15.0 (2.0 - 161.0) | 29.0 (38.2) | 43.0 (2.0 - 161.0) | 6.0 (9.9) | 7.0 (2.0 - 132.0) |

The table divides the study population into a group with all enrolled patients (“Overall”) and a group stratified by birth weight categories of < 1,500 g and ≥ 1,500 g.

#### eTable 4: WGS-Cluster-Contributing MDRO+

| MDRO+ | ST | WGS Cluster | Number of Patients involved |
|---|---|---|---|
| *A. baumannii* | fusA_SLV_ST506 | 202006_fusA_SLV_ST506 | 3 |
| *A. baumannii* | C | 201906_C | 2 |
| *E. cloacae* | 106 | 201903_ST106 | 2 |
| *E. cloacae* | 114 | 202006_ST114 | 5 |
| *E. cloacae* | 32 | 202001_ST32 | 4 |
| *E. cloacae* | 50 | 201911_ST50 | 2 |
| *E. cloacae* | 61 | 201904_ST61 | 3 |
| *E. cloacae* | 68 | 201907_ST68 | 5 |
| *E. cloacae* | 78 | 202002_ST78 | 3 |
| *E. cloacae* | 0Q | 201907_ST0Q | 3 |
| *E. cloacae* | 0AK | 202005_ST0AK | 4 |
| *E. coli* | 1331 | 202008_ST1331 | 8 |
| *E. coli* | 141 | 201903_ST141 | 3 |
| *E. coli* | 141 | 201905_ST141 | 15 |
| *E. coli* | 141 | 201908_ST141 | 10 |
| *E. coli* | 404 | 201903_ST404 | 6 |
| *E. coli* | 405 | 201903_ST405 | 2 |
| *E. coli* | 420 | 201910_ST420 | 4 |
| *E. coli* | 73 | 201902_ST73 | 8 |
| *E. coli* | 73 | 201907_ST73 | 3 |
| *E. coli* | 73 | 201911_ST73 | 5 |
| *E. coli* | 83 | 201903_ST83 | 2 |
| *K. oxytoca* | 146 | 202002_ST146 | 8 |
| *K. oxytoca* | 151 | 202005_ST151 | 2 |
| *K. oxytoca* | 176 | 201903_ST176 | 1 |
| *K. oxytoca* | 176 | 202006_ST176 | 19 |
| *K. oxytoca* | 186 | 202005_ST186 | 4 |
| *K. oxytoca* | 40 | 201911_ST40 | 2 |
| *K. oxytoca* | 48 | 201909_ST48 | 2 |
| *K. pneumoniae* | 1486 | 201909_ST1486 | 2 |
| *K. pneumoniae* | 20 | 202008_ST20 | 3 |
| *K. pneumoniae* | 219 | 201908_ST219 | 6 |
| *K. pneumoniae* | 2974 | 201908_ST2974 | 6 |
| *K. pneumoniae* | 966 | 202006_ST966 | 7 |
| MSSA | 22 | 202002_ST22 | 3 |
| MSSA | 5 | 201907_ST5 | 2 |
| MSSA | 8 | 201910_ST8 | 3 |
| *S. marcescens* | NA | 202001 | 7 |
| Total | | | 179 |

MDRO+ species with respective ST, WGS-cluster-definition and numbers of patients involved (including index patients).

#### eTable 5: WGS-non-Cluster-Contributing MDRO+ ("singletons")

| MDRO+ | ST | WGS Cluster | Number of Patients involved |
|---|---|---|---|
| *A. baumannii* | 106 | singleton | 1 |
| *A. baumannii* | 205 | singleton | 1 |
| *A. baumannii* | 93 | singleton | 2 |
| *A. baumannii* | 1459 | singleton | 1 |
| *C. freundii* | 162 | singleton | 1 |
| *C. freundii* | 64 | singleton | 1 |
| *C. freundii* | 98 | singleton | 1 |
| *C. freundii* | NA | singleton | 2 |
| *E. cloacae* | 1283 | singleton | 1 |
| *E. cloacae* | 145 | singleton | 5 |
| *E. cloacae* | 158 | singleton | 1 |
| *E. cloacae* | 278 | singleton | 1 |
| *E. cloacae* | 286 | singleton | 2 |
| *E. cloacae* | 664 | singleton | 1 |
| *E. cloacae* | 672 | singleton | 1 |
| *E. cloacae* | 78 | singleton | 1 |
| *E. cloacae* | 831 | singleton | 1 |
| *E. cloacae* | NA | singleton | 3 |
| *E. coli* | 10 | singleton | 2 |
| *E. coli* | 101 | singleton | 1 |
| *E. coli* | 1154 | singleton | 1 |
| *E. coli* | 117 | singleton | 1 |
| *E. coli* | 1193 | singleton | 1 |
| *E. coli* | 127 | singleton | 2 |
| *E. coli* | 131 | singleton | 1 |
| *E. coli* | 1314 | singleton | 1 |
| *E. coli* | 14 | singleton | 1 |
| *E. coli* | 144 | singleton | 1 |
| *E. coli* | 2015 | singleton | 1 |
| *E. coli* | 2166 | singleton | 1 |
| *E. coli* | 23 | singleton | 1 |
| *E. coli* | 295 | singleton | 1 |
| *E. coli* | 38 | singleton | 1 |
| *E. coli* | 399 | singleton | 1 |
| *E. coli* | 3LV | singleton | 1 |
| *E. coli* | 404 | singleton | 1 |
| *E. coli* | 405 | singleton | 2 |
| *E. coli* | 428 | singleton | 2 |
| *E. coli* | 569 | singleton | 1 |
| *E. coli* | 5910 | singleton | 1 |
| *E. coli* | 62 | singleton | 1 |
| *E. coli* | 657 | singleton | 1 |

| | | | |
|---|---|---|---|
| *E. coli* | 69 | singleton | 5 |
| *E. coli* | 6949 | singleton | 1 |
| *E. coli* | 73 | singleton | 1 |
| *E. coli* | 80 | singleton | 2 |
| *E. coli* | 95 | singleton | 2 |
| *E. coli* | 967 | singleton | 1 |
| *E. coli* | icd_SLV_ST73 | singleton | 1 |
| *K. aerogenes* | 135 | singleton | 1 |
| *K. aerogenes* | NA | singleton | 3 |
| *K. oxytoca* | 11 | singleton | 1 |
| *K. oxytoca* | 13 | singleton | 1 |
| *K. oxytoca* | 176 | singleton | 1 |
| *K. oxytoca* | 178 | singleton | 1 |
| *K. oxytoca* | 184 | singleton | 1 |
| *K. oxytoca* | 36 | singleton | 1 |
| *K. oxytoca* | 40 | singleton | 2 |
| *K. oxytoca* | 64 | singleton | 1 |
| *K. oxytoca* | 84 | singleton | 1 |
| *K. oxytoca* | 88 | singleton | 1 |
| *K. oxytoca* | gapA_SLV_ST111 | singleton | 1 |
| *K. oxytoca* | pgi_phoE_DLV_ST157 | singleton | 1 |
| *K. oxytoca* | NA | singleton | 3 |
| *K. pneumoniae* | 105 | singleton | 1 |
| *K. pneumoniae* | 13 | singleton | 1 |
| *K. pneumoniae* | 1731 | singleton | 1 |
| *K. pneumoniae* | 1875 | singleton | 1 |
| *K. pneumoniae* | 1962 | singleton | 1 |
| *K. pneumoniae* | 200 | singleton | 2 |
| *K. pneumoniae* | 2478 | singleton | 1 |
| *K. pneumoniae* | 26 | singleton | 1 |
| *K. pneumoniae* | 292 | singleton | 1 |
| *K. pneumoniae* | 308 | singleton | 1 |
| *K. pneumoniae* | 397 | singleton | 2 |
| *K. pneumoniae* | 433 | singleton | 1 |
| *K. pneumoniae* | 45 | singleton | 2 |
| *K. pneumoniae* | 465 | singleton | 1 |
| *K. pneumoniae* | 48 | singleton | 1 |
| *K. pneumoniae* | 596 | singleton | 1 |
| *K. pneumoniae* | 792 | singleton | 1 |
| *K. pneumoniae* | 883 | singleton | 1 |
| *K. pneumoniae* | NA | singleton | 2 |
| MSSA | 1 | singleton | 2 |
| MSSA | 15 | singleton | 4 |
| MSSA | 22 | singleton | 2 |
| MSSA | 30 | singleton | 3 |

| | | | |
|---|---|---|---|
| MSSA | 398 | singleton | 5 |
| MSSA | 45 | singleton | 2 |
| MSSA | 46 | singleton | 1 |
| MSSA | 5 | singleton | 1 |
| MSSA | 582 | singleton | 3 |
| MSSA | 672 | singleton | 1 |
| MSSA | 7 | singleton | 3 |
| MSSA | 72 | singleton | 1 |
| MSSA | 8 | singleton | 2 |
| MSSA | 938 | singleton | 1 |
| MSSA | 946 | singleton | 2 |
| MSSA | NA | singleton | 3 |
| *P. aeruginosa* | 108 | singleton | 1 |
| *P. aeruginosa* | 395 | singleton | 2 |
| *S. marcescens* | NA | singleton | 7 |
| Total | | | 157 |

MDRO+ species with respective ST, WGS-cluster-definition and numbers of patients involved.

#### eTable 6: AFLP-Mismatches for *E. coli*

| *E. coli* | AFLP-Type Matching WGS-Cluster | | AFLP-Type does not match WGS-Cluster | |
|---|---|---|---|---|
| Clusters | AFLP Type | Count | AFLP Type | Count |
| 201902_ST73 | ***E*** | 8 | - | - |
| 201907_ST73 | - | - | ***E*** | 3 |
| 201911_ST73 | AL | 5 | - | - |
| 201903_ST404 | P | 6 | - | - |
| 201903_ST405 | ***AX*** | 2 | ***AW*** | 1 |
| 201903_ST83 | C | 2 | - | - |
| 201905_ST141 | ***D*** | 15 | - | - |
| 201908_ST141 | - | - | ***D*** | 9 |
| 201903_ST141 | - | - | ***D*** | 3 |
| 201910_ST420 | ***AH*** | 2 | ***AY*** | 1 |
| 202008_ST1331 | K | 6 | - | - |

Detailed allocation of AFLP-types to WGS-clusters for *E. coli*. Mismatched AFLP-Types in bold/cursive. AFLP-Types “D” and “E” successfully identified ST141 and ST73 respectively. WGS cluster definitions enabled further separation of *E. coli* Sequence Types. For example ST 141 can be separated into distinct WGS clusters “201903_ST141” [3], “201905_ST141 [15], “201908_ST141 [10]”.

#### eTable 7: Cumulative AFLP-Mismatch Rates per MDRO+

| MDRO+ | Cumulative Mismatch Rate (AFLP vs. WGS) | Number of WGS Clusters |
|---|---|---|
| *A. baumannii* | 0,00 | 2 |
| *E. cloacae* | 0,09 | 9 |
| *E. coli* | 0,06 | 11 |
| *K. oxytoca* | 0,10 | 6 |
| *K. pneumoniae* | 0,08 | 5 |
| *S. marcescens* | 0,00 | 1 |
| MSSA | 0,11 | 3 |

Based on the less granular resolution of AFLP-typing in comparison to WGS we observed cumulative mismatch rates for respective MDRO+ species. Note: S. *aureus* underwent *spa*-typing as AFLP-typing is inadequate for this species.

### eTable 8: WGS Enabled Net Gain in Resolution

| Cluster (Species Level) | Cluster Start | Cluster End | WGS Cluster with Overlapping Timeframe of Species Cluster | Gain in Resolution |
|---|---|---|---|---|
| Escherichia_coli_1 | 2019-02-21 | 2020-03-18 | 201903_ST141, 201903_ST83, 201902_ST73, 201903_ST404, 201905_ST141, 201907_ST73, 201908_ST141, 201910_ST420, 201911_ST73, 201903_ST405 | 9 |
| Klebsiella_oxytoca_12 | 2020-05-16 | 2020-10-28 | 202005_ST151, 202005_ST186, 201911_ST40, 202006_ST176, 201903_ST176 | 4 |
| Staphylococcus_aureus_3 | 2019-10-14 | 2020-03-09 | 201910_ST8, , 202002_ST22 | 2 |
| Enterobacter_cloacae_4 | 2019-06-16 | 2019-09-09 | 201907_ST68, 201907_ST0Q, | 2 |
| Klebsiella_pneumoniae_7 | 2019-08-25 | 2020-02-28 | 201908_ST219, 201908_ST2974, 201909_ST1486 | 2 |
| Enterobacter_cloacae_8 | 2020-05-10 | 2020-07-03 | 202005_ST0AK, , 202006_ST114 | 2 |
| Enterobacter_cloacae_1 | 2019-02-24 | 2019-03-14 | 201903_ST106, | 1 |
| Staphylococcus_aureus_1 | 2019-02-17 | 2019-09-11 | 201907_ST5, | 1 |
| Klebsiella_pneumoniae_10 | 2020-05-10 | 2020-10-29 | 202006_ST966, 202008_ST20 | 1 |
| Acinetobacter_baumannii_1 | 2019-06-10 | 2019-08-02 | 201906_C | 0 |
| Klebsiella_oxytoca_5 | 2019-09-05 | 2019-09-17 | 201909_ST48 | 0 |
| Acinetobacter_baumannii_7 | 2020-06-01 | 2020-08-07 | 202006_fusA_SLV_ST506 | 0 |
| Escherichia_coli_10 | 2020-08-10 | 2020-11-01 | 202008_ST1331 | 0 |
| Klebsiella_oxytoca_11 | 2020-02-16 | 2020-04-28 | 202002_ST146 | 0 |

Table of species-level clusters that overlap with WGS clusters in time and the gain in cluster-resolution. The day of detection of an MDRO+ defines the start date. The discharge of the last patient to be colonized with respective cluster MDRO+ defines the end date. This logic applies for both clusters on species and WGS level.
